# “This guideline is not practical; it might be practical to specialists like you”: Doctors’ experiences of the Global Initiative for Asthma (GINA) guideline in Nigeria; A qualitative study

**DOI:** 10.1101/2022.10.14.22281114

**Authors:** Obianuju Beatrice Ozoh, Sandra Kelechi Dede, Joy Nkiru Eze, Kevin Mortimer, Martha Chinouya

## Abstract

**Background:** The Global Initiative for Asthma (GINA) report sets out updated evidence-based strategy for asthma management. Little is known about how this report is perceived and implemented in low-income and middle-income countries (LMICs) like Nigeria. We explored the experiences of asthma management as informed by the current GINA guideline among doctors in Lagos, Nigeria.

**Methods:** Using a qualitative research approach, in-depth interviews were conducted among doctors in Lagos, Nigeria to explore their experiences of asthma management in the context of the current GINA report. The thematic framework approach was used for data analysis.

**Results:** Eleven doctors (five general practitioners (GPs) and six Family Physicians (FPs) took part. Four overarching themes were identified:

1. Knowledge of, and attitude towards the GINA strategy: Whilst most doctors were aware of the existence of the GINA report, there was limited knowledge about its content including current recommendations for mild asthma treatment.
2. Asthma diagnosis and treatment: There was limited access to lung function testing facilities and its role in asthma diagnosis was underappreciated.
3. Barriers to managing asthma according to GINA recommendations: These included complexity of the GINA report, unavailability and unaffordability of asthma medicines and poor patient adherence to medications, driven by socio-cultural factors.
4. Enablers of GINA-recommended asthma management: Improvement in asthma education for doctors and the general population and better access to diagnostic tests and medicines.

**Conclusion:** Whilst there was awareness of the existence of the GINA report, there was limited knowledge about its content and several barriers to its implementation were reported. Education about the GINA report, investment in diagnostic and treatment services and simplification of recommendations were identified as possible solutions.

## Introduction

Asthma is the most common chronic respiratory disease (CRD) in children and remains one of the most common CRDs through adolescence and into adulthood [1–4]. The great majority of the 260 million people with asthma globally live in low-income and middle-income countries (LMICs) where they face a disproportionately high burden of morbidity (84% of global disability-adjusted life years) and mortality (96% of global asthma related deaths) [5,6]. In Nigeria, a LMIC in sub-Saharan Africa (sSA) with a population of around 200 million people, the prevalence of asthma is 6.2%.[7] Fewer than 10% of these have well controlled disease suggesting that asthma is a major cause of avoidable morbidity and potentially mortality in Nigeria and other LMICs [6,8,9].

There are many challenges to asthma management in LMIC. These range from limited availability and affordability of asthma medicines and diagnostics to low prioritization of asthma by policy makers [10,11,12]. Many LMIC lack National strategies and updated essential medicine lists for asthma management [13].

The Global Initiative for Asthma (GINA) publishes an annually updated evidence-based strategy for asthma. In 2019, an important change to GINA recommendations was made such that inhaled corticosteroids (ICS) are now recommended for everyone with asthma, either just as and when an inhaled bronchodilator is taken or as maintenance treatment and as required depending on the frequency of symptoms and risk of exacerbations [14,15]. The rationale for this change was the underpinning evidence that persons with mild asthma could have serious, life threatening and fatal exacerbations and are usually poorly adherent to daily ICS [15]. The Nigerian asthma guidelines have not yet been updated in the same way [16].

The GINA strategy can be adapted for use in individual countries in consideration of national health systems and availability and affordability of asthma diagnostic and treatment services [14]. Over recent years, GINA has been working to maximize the relevance of the GINA report to LMICs for example through recommendations in the report that address resource-limited settings and publications that put the GINA report in the context of the reality of asthma management in LMICs [9,17]. However, little is known about how effective these efforts have been or how the GINA report is perceived and implemented in LMICs like Nigeria. To help provide some insights into this, we did a qualitative study exploring the experiences of doctors in Lagos, Nigeria of asthma management in the context of the current GINA report.

## Materials and Methods

### Design

This was a qualitative study in which in-depth interviews were conducted using a topic guide (Appendix 1). The topic guide was iteratively developed and informed by the literature and experience of the first author, a Nigerian pulmonologist and lecturer who also manages asthma at a tertiary care hospital in Lagos.

### Setting

Nigeria is the most populous African country and over 40% of the population live below the poverty line [18]. Lagos is the most cosmopolitan and populated city in Nigeria, with high levels of air pollution and the highest prevalence of asthma [7]. The healthcare system in Nigeria is poorly resourced and biomedical healthcare is provided through public (67%) and private health facilities (33%) at the primary (88%), secondary (12%) and tertiary levels (0.25%), with 76% of secondary facilities being privately owned [19,20]. About 90% of patients bypass the primary healthcare facilities due to perceived need for quality services and self-refer to the secondary care facilities [21]. Doctors provide services at secondary facilities with a greater use of task shifting at primary care centres. Most general practice clinicians which comprise General Practitioners (GPs), with only undergraduate medical training, and Family Physicians (FP), who have postgraduate residency training, work at secondary care facilities and private hospitals, where most asthma patients seek care [8].

### Participants/ Sampling approach

Participants comprised FPs and GPs who worked at public and private hospitals. They were included if they attended to a minimum of three asthma patients in a month. Doctors who worked at tertiary hospitals and house officers were excluded.

Purposive sampling was used to select doctors who had personal experience of asthma management. We aimed for near equal distribution between GPs and FPs as well as between publicly funded and private hospitals.

Participants were identified through a network of professional colleagues and by snowballing and contact established by email. Those who consented to participate were recruited and a date set for interview. No monetary compensation was provided.

We aimed to recruit 10-15 participants and stopped when we achieved saturation of themes.

### Data Collection

Data collection took place between April and May 2021. The interviews were conducted in a Coronavirus-2019 (COVID-19) secure manner using video teleconferencing technology (Zoom®) with participants alone in a quiet room.[22]

Semi-structured interviews were conducted by the first author. Open-ended questions and probes which allowed open discussion and promoted the emergence of new ideas to enrich the data were used.[23] Interviews were conducted in English which is spoken fluently by all doctors and lasted 45-60 minutes. Participants’ demographics, duration of practice, hospital setting and availability of asthma diagnostic tools (spirometry and peak flow meter) at their hospital were obtained.

### Data Management

Interviews were recorded and saved on a secure password-protected computer prior to transcription. Recordings were transcribed using the denaturalized approach.[24] Fields notes and a reflexive log taken during interview were used to annotate areas where there was a threat to confirmability.

### Data Analysis

We used the framework approach and iteratively applied codes which were then categorized to develop a thematic framework.[25],[26],[27]. Indexing was done across all text and charted across themes in Nvivo® (QRS International, Melbourne, Australia). Data were mapped and interpreted by identifying associations, providing explanations and creating typologies.

## Results

We recruited 11 doctors (age 28-46 years; 6 females; five FPs and six GPs) (Table 1). Spirometry was available at three private hospitals and a peak flow meter was available at one private hospital without spirometry.

**Table 1:** Characteristics of the study participants and their practice setting.

| Characteristic | Description |
| --- | --- |
| Age range in years | 28-46 |
| Sex |  |
| Male | 5 |
| Female | 6 |
| Level of qualification |  |
| General practitioner | 6 |
| Family physician | 5 |
| Duration of practice (range) in years | 5-21 years |
| Public funded hospitals | 5* |
| Private hospitals | 6 |
| Availability of spirometer |  |
| Private hospital | 3 |
| Public hospital | 0 |
| Availability of the peak flow |  |
| Private hospital | 1 |
| Public hospital | 0 |
| Awareness of the GINA 2019 update | 3** |
| Awareness of the Nigerian asthma guideline | 4 |

Footnote: \* Two participants were from the same hospital. \*\*All participants aware of the GINA update were family physicians.

Three doctors (all FPs) were aware of the new GINA recommendation for the treatment of mild asthma. Two FPs and two GPs were aware of the existence of the Nigerian asthma guideline but had never read it.

Four overarching themes were identified and are described below, buttressed by quotes from the participants.

These were 1). Knowledge of and attitude towards the GINA strategy. 2). Asthma diagnosis and treatment. 3). Barriers to managing asthma according to GINA recommendations. 4). Enablers of GINA-recommended asthma management.

### 1. Knowledge of and Attitude towards the GINA Strategy

Most participants were aware of but had never read the GINA guideline and therefore had poor knowledge of its content. Knowledge was mostly obtained during medical school and or at CME programmes. The GPs considered it unnecessary to know the details of the strategy as one GP noted:

> *“I’m aware of the GINA guideline but to be frank, I’ve never really been able to know the details. I just know that there is the GINA guideline. I have been managing it the way we’ve been doing it right from housejob and the simple way we do it as medical officers in my centre of practice now…. I am very sure I am not managing according to the guidelines”. The maximum that I do in my practice is just to relieve patients*. (GP, private hospital)

There was also low awareness of the Nigerian asthma guideline. It was considered inaccessible compared to international guidelines. A GP who was aware of the Nigerian asthma guideline put it this way:

> *“Well, lately it’s been more of the UK version actually, I find it quite clear in the steps… the, Nigerian version is dated …, and it is not as clear. The NICE one has very obvious steps along the way, what to do…. when you routinely search for guidelines about a specific condition I think, the US and UK bodies have easier access on google to prop up”*. (GP, private hospital)

Despite the low levels of awareness of the content of GINA, there was a positive attitude towards guidelines generally as participants considered guidelines trustworthy, evidence-based and capable of promoting standardised practice. However, most participants had reservations towards the suitability and applicability to local contexts such as Lagos. This is how one FP opined when discussing how guidelines are often developed in high income countries with the appropriate diagnostic resources:

> *“…but I think the problem that we have most times especially those of us back here are that sometimes most of these guidelines are done with the Caucasians, so sometimes what suits them doesn’t suit us”*. (FP, public hospital).

Importantly, guidelines were not considered to be rules with legal recourse; therefore, adherence to recommendations from the guidelines in their practices, was therefore unprioritized. They reported that observation of senior colleagues or hospital norms regarding treatment usually informed their practice and saved time. New information from CMEs could also influence practice, but these were usually dependent on the availability of resources for implementation. One GP reported how management of asthma is experiential and shared within their practice:

> *“So, it’s not easy to always go to the guidelines. There are many things to do, many patients to see. There is a standard way we know that has been passed down from one medical officer to another…. We just adopt the way senior colleagues in the job have been doing it and we just continue that way”*. (GP, private hospital).

There was a positive attitude towards the GINA recommendation for treatment with ICS for all severities of asthma, but, they questioned the feasibility due to unavailability and unaffordability of ICS:

> *“For GINA to come up with this, that means that they have done their research …and found that it is best for asthma patients to be on continuous steroids. So, I see that as good*… *“I don’t see it as feasible in our environment, mainly because of lack of access and the cost, even availability, how available is it”?* (FP, private hospital).

### 2. Asthma Diagnosis and Treatment

For most participants, asthma diagnosis did not involve lung function testing using spirometry or the peak flow. Most were dispassionate about the GINA recommendation for lung function testing and felt it was of no value. The three doctors with access to spirometry did not use it because it was impracticable during exacerbations when most patients presented. One FP who has access to spirometry said:

> *“So, that’s how I make the diagnosis, mainly using the symptoms…. Well, there is really no reason why I don’t consider it. It’s just that I don’t, I have not really been, I am just not keen about telling them to do the test…. for me to see a patient in an acute state and begin to do, ask the patient, go and do tests, for me it is delay, so treat the patient immediately even if it is symptomatically*.*”* (FP, private hospital)

The only doctor with access to peak flow however found it valuable and she used it regularly. She said:

> *“So, I just used that example…. to give you an idea of how just using a basic peak flow meter, even in the absence of spirometry, it was able to help me make a diagnosis…decision of whether it is asthma or something else*.*”* (FP, private hospital)

GPs perceived asthma as an acute disease and reported that the focus in their practice was to avoid triggers for asthma flare ups and treatment of exacerbations. However, FPs were more likely to consider asthma as a chronic disease as they regularly saw individual patients presenting in their practices over time. Subsequently, GPs reported that they had ‘low confidence’ in making long term decisions for asthma treatment and deferred to specialists. This GP describes:

> *“…And I am very sure I am not managing according to the guidelines…After they are relieved and stable to go home on salbutamol inhalers, we refer them to the pulmonology clinic for long term care… It’s the specialist that does the real diagnosis*…*”* (GP, private hospital).

When discussing treatment plans for their patients including training patients on inhaler techniques, most participants did not mention reference to the guideline recommendations. Due to time constraints, GPs reported that in most cases, they referred patients to pharmacists for the training. Only one FP shared experience with the use of written asthma action plan and none objectively assessed asthma control. This GP reports:

> *“Because of time constraints or because of doctor even being tired because sometimes you can actually see up to 50 patients…”. Nobody taught them how to use the inhaler…and the pharmacist might not be able to teach or might not even have the time …”* (FP, public hospital).

The participants reported that, in their practices, they had experienced patient inhaler hesitancy and non-adherence to treatment. They attributed these patient factors to cultural beliefs and myths about asthma. As captured in this quote:

> *“Like maybe from 60% do not tend to initially accept. Because they have like, will I say, wrong notion that when they use the inhaler, the diagnosis is like permanent or how do I put it-they now really get the asthma. They don’t understand that the inhaler is to help, to relieve symptoms. There are some that will rather prefer to take oral medications than to take an inhaler”*. (GP, public hospital)

One participant described how patients preferred to use complementary and alternative medicines and said:

> *“* .. *a local stone they throw in hot water and inhale the steam …*.*they are quick to tell you that this is what they have tried and this what has happened … And interestingly even though it doesn’t work negative reviews and criticism do not follow”*. (GP, private hospital)

Another GP describes how patients’ experience of stigma influenced adherence:

> *“Most people will not want to be seen with this inhaler*… *It will come with a bad attitude or rather a bad perception of “are you sick? What is going on?” and it is definitely going to reduce the likelihood of using it… and letting people know that you even have asthma…. Definitely, there are a lot of cultural bias in there”*. (GP, public)

Participants also reported how spirituality influenced doctors’ experiences in diagnosing and treating asthma. They all admitted to having spiritual beliefs which influenced their practice when encountering patients with asthma:

> *I am a Christian, that helps me …*.*to listen to their complaints, their fears…so I now use that aspect of spirituality as a form of comfort to them while I still go ahead to give them the proper treatment”* (FP, private hospital).

She also added how patients’ own spirituality influences their responses to the diagnosis of asthma:

> *“*..*when you tell them that this is likely asthma, they say, “ No, God forbid, it is not me, I can’t have asthma”,… “my enemy has asthma it is not me”…*.*Nigerians are very spiritual people…There’s a big clash between spirituality and medical care*.
>
> *So, it takes extra effort and I tend to spend enough time…trying to counsel them… …Based on my own view, I’ve learnt to separate medicine and religion*…

The participants reported that they had experienced poor attendance for follow up care which hinged on the intermittent nature of asthma, poor health seeking behaviour and difficulty accessing care at public hospitals. One FP said:

> *“You know that in Nigeria people don’t like going to hospital, so they already have an idea of how to manage themselves and they don’t come unless it is that bad*.*”* (FP, public)

#### Another GP said

> *“they don’t see the need or the essence or the importance of coming back for check-up and maybe on our part we need to do more on that like waiting time to see a doctor”*. (GP, public)

### 3. Barriers to Managing Asthma according to GINA Recommendations

The GINA report was considered, by the participants, as cumbersome and impractical due to the multiple treatment steps. The Nigerian asthma guideline was not expected to provide additional value as it was opined to likely be a copy of the GINA recommendations without consideration of the local context. One description of the GINA guideline was:

> *“The GINA guideline is not something that is practical, it might be practical to specialists like you (referring to the interviewer) but for ordinary doctors like us, we don’t really see it as being practical”*. (FP, private)

The doctors also reported that the unavailability and unaffordability ICS hindered access and use. Doctors in public hospitals experienced stock-out of ICS inhalers more regularly. A GP said:

> *“But if it is expensive or not easily available…they start looking for a cheaper alternative, which is the Ventolin that is everywhere and they can just pick up”*. (GP, public).

A FP also noted that insurance coverage for ICS was limited which further hindered access due to costs:

> *“99% of the patients we see are HMOs’ and most times they will not give them “Seretide” because it’s expensive”*. (FP, public)

### 4. Enablers of GINA-recommended Asthma Management

Participants also discussed ways of enabling doctors working within the context to implement GINA recommendations. A universally recognized enabler was delivering more CMEs for general practice clinicians as they considered themselves as frontline workers. One GP felt that that asthma education was low compared to other chronic diseases, which they could learn from, and said:

> *“I know that some companies are actually doing that for diabetes because recently I just filled a form for…International Diabetic Federation for training on diabetes trying to equip the health care professionals on the best practice in managing diabetes. I believe if we can do the same for asthma, we can have more practitioners having a good grasp on managing asthma”*. (GP, public)

There was also a strong recommendation for asthma education of the general public to improve population health literacy on the condition. They believed, at a population level, education would reduce the negative influence of myths, fight stigma associated with asthma, with the perceived outcome of improving support and adherence to medication. This FP put it this way:

> *“Increased awareness of asthma through the media. I don’t think I have seen anything. I see them talk about hypertension, I see them talk about TB but I don’t think I have really seen much media publicity about asthma…”* (FP, private).

Clinicians laid responsibility on the government for multiple aspects of asthma care such as, improving the availability and affordability of asthma medicines through Universal Health Coverage, reducing air pollution and regular practice audits. They suggested:

> *“I know asthma is not necessarily avoidable but… if the environment is cleaner, there are less fumes, pollutants and things like that, that might to an extent help”*. (FP, public).

#### Others added

> *“…in other parts of the world reviews of doctors (is) a continuous effort …there should be uniformity in their thoughts and the way they manage because there is a body that actually penalizes and ensures that doctors are following through…but not in Nigeria…if you ask this person why do you do this? This Is how I have been doing it”*. (GP, private)

## Discussion

This is the first qualitative study to explore the experience of doctors in Nigeria in using the GINA guidelines in asthma management. This study adds to the ongoing debates regarding adherence to guidelines in the management of chronic respiratory diseases. We found that whilst there was widespread awareness of the existence of the GINA strategy amongst the doctors in Nigeria included in this study, they had poor knowledge of its content, including the current treatment update. The FPs had better knowledge compared to the GPs. Despite positive attitude towards guidelines, asthma diagnosis and treatment were informed by hospital norms rather than the GINA or the national asthma guideline. Echoing findings from previous reports on access to lung function test equipment, the participants reported that there was limited access to lung function testing facilities, but added that it was mostly undervalued for asthma diagnosis [11,28]. The doctors found it reasonable and were willing to use ICS for all asthma patients but the feasibility of domesticating it in practice was uncertain due to limited availability and unaffordability of the medications. They experienced inhaler hesitancy and poor adherence to treatment among patients, driven by myths, cultural practices, stigma, spirituality, and preference for alternative medicines. Barriers to guideline-based management included complexity of the guideline and inaccessibility of the facilities and medicines for treatment. Enablers proffered included improvement in asthma education and access to facilities for management.

Low use of lung function testing increases the risk of both under- and over-diagnosis of asthma.[29] Although limited access to spirometry equipment in Nigeria is recognized, attitudes of doctors towards this test affects utilization [30]. This implies that improvement in access to spirometry alone therefore may not impact asthma management. Peak flow measurement is favoured in many primary care services even where there was access to spirometry because of its simplicity and the immediacy of the results [31]. Considering the much lower resource need in using the peak flow, it is recommended for first line lung function testing for general practice doctors. This recommendation is supported by GINA and included in the World Health Organization (WHO) Package of essential non communicable (PEN) disease interventions catalog as an essential tool in the management of CRD [32,33].

Despite the clear benefits of the use of ICS across all asthma severities, most doctors doubted the feasibility. Unavailability and unaffordability of the medicines for the patients, coupled with lack of UHC for asthma treatment were the main reasons. Medication costs account for the highest out of pocket expenditure in asthma treatment and contributes to poor patient adherence [34]. Central medication procurement and UHC for controller medications are approaches that could improve access [28]. Medical syncretism which is pervasive in Nigeria and other parts of Africa especially for chronic diseases such as asthma and also contributes to poor adherence [35]. Stigma including self-stigmatization was also an important hinderance to guideline-based asthma management, promoting non-adherence to treatment. These are important considerations for asthma education in Nigeria and other African countries where health literacy is low and cultural factors strongly influence health behaviours [35]. Improving population literacy in these settings is of high priority.

The structure of the GINA strategy, which was described as cumbersome, was another barrier to use. Power in the context of guideline development was raised by participants and inclusion is needed in this regard. However, contextualization of local asthma guidelines such as the Nigerian asthma guideline by simplifying and streamlining treatment recommendations to locally available and affordable medicines could address this.

The reliance of doctors on hospital norms for asthma treatment, their poor attitude towards guideline recommended practice and the inadequacy of facilities for optimal patient care is arguably structural violence, perpetuating poor patient care [36]. Based on the experiences of these participants and previous reports, many African health systems are focused on the delivery of emergency asthma treatment [37]. There is a strong need to prioritize the care of patients with CRD such as asthma in many parts of Africa as a matter of equity and justice.

We acknowledge that our study had limitations. Restricting participants to GPs and FPs excluded some clinicians who treat asthma although most asthma patients in Nigeria see this category of doctors. A potential weakness of in-depth interviews is the risk of the Hawthorne effect, however by being self-aware and reflexive, the interviewer reduced hierarchical power and promoted open and honest discussion. Through iterative questioning and member checking during the interview we improved the confirmability of our findings. The limitations of virtual interviews with regards to establishing rapport and appreciating the setting were partly mitigated by the use of video [38,39].

Our study also had strengths. To our knowledge this is the first qualitative study that has explored factors explaining poor adherence to GINA recommendations among Nigerian doctors. The research team of local clinicians and experienced qualitative researchers enhanced the credibility of our findings. There was regular debriefing and reflection during data collection promoting objectivity.

## Conclusions

Whilst there was awareness of the existence of the GINA report, there was limited knowledge about its content amongst the doctors who took part in this study in hospitals in Lagos, Nigeria. Several barriers to the implementation of GINA recommendations into practice were identified including limited access to diagnostic tests (particularly spirometry), reluctance to prescribe inhaled treatments, poor treatment concordance and limited access to affordable asthma medicines. Education based on the GINA report, investment in diagnostic and treatment services and simplification of asthma management recommendations are possible solutions.

## Data Availability

Data will be available from the corresponding author on reasonable request.

## Ethics and Consent Statement

Ethical approval was obtained from the Masters Research Ethics Committee (MREC) of Liverpool School of Tropical Medicine (LSTM) (APP No.:GH21(01)); and the Lagos University Teaching Hospital (LUTH), Lagos, Nigeria (Assigned No.: ADM/DCST/HREC/APP/4046). All participants provided written informed consent.

## Authors’ contribution

**Conceptualization**: Obianuju B. Ozoh

**Data Curation**: Obianuju B. Ozoh

**Formal Analysis**: Obianuju B. Ozoh, Sandra K. Dede

**Funding acquisition**: Not applicable

**Methodology**: Obianuju B. Ozoh, Sandra K. Dede, Joy N. Eze, Martha Chinouya

**Project administration**: Obianuju B. Ozoh, Martha Chinouya, Kevin Mortimer

**Supervision**: Martha Chinouya, Kevin Mortimer

**Writing- original draft**: Obianuju B. Ozoh, Sandra K. Dede, Joy N. Eze, Kevin Mortimer Martha Chinouya.

**Writing-review and editing**: Obianuju B. Ozoh, Sandra K. Dede, Joy N. Eze, Kevin Mortimer Martha Chinouya.

## Acknowledgements

We acknowledge the family physicians and the general practitioners for their cooperation in all phases of the data collection.

## Abbreviations

CRD: Chronic respiratory diseases
DALYs: Disability Adjusted Life Years
FPs: Family physicians
GPs: General Practitioners
GINA: Global Initiative for Asthma
ICS: Inhaled corticosteroids
LMIC: Low-income and Middle-Income Countries
NCD: Non-communicable disease

## References

1. Wang H, Naghavi M, Allen C, Barber RM, Bhutta ZA, Carter A, et al. Global, regional, and national life expectancy, all-cause mortality, and cause-specific mortality for 249 causes of death, 1980–2015: a systematic analysis for the Global Burden of Disease Study 2015. The Lancet. 2016;388: 1459–1544. doi:10.1016/S0140-6736(16)31012-1

2. Mortimer K, Nantanda R, Meghji J, Vanker A, Bush A, Ndimande N, et al. Africa’s respiratory “Big Five.” JPATS. 2021;2: 64–72. doi:10.25259/JPATS_12_2021

3. Mortimer K, Lesosky M, García-Marcos L, Innes Asher M, Pearce N, Ellwood E, et al. The burden of asthma, hay fever and eczema in adults in 17 countries: GAN Phase I study. Eur Respir J. 2022; 2102865. doi:10.1183/13993003.02865-2021

4. García-Marcos L, Innes Asher M, Pearce N, Ellwood E, Bissell K, Chiang C, et al. The burden of asthma, hay fever and eczema in children in 25 countries: GAN Phase I study. Eur Respir J. 2022; 2102866–2102866. doi:10.1183/13993003.02866-2021

5. Soriano JB, Abajobir AA, Abate KH, Abera SF, Agrawal A, Ahmed MB, et al. Global, regional, and national deaths, prevalence, disability-adjusted life years, and years lived with disability for chronic obstructive pulmonary disease and asthma, 1990–2015: a systematic analysis for the Global Burden of Disease Study 2015. The Lancet Respiratory Medicine. 2017;5: 691–706. doi:10.1016/S2213-2600(17)30293-X

6. Meghji J, Mortimer K, Agusti A, Allwood BW, Asher I, Bateman ED, et al. Improving lung health in low-income and middle-income countries: from challenges to solutions. The Lancet. 2021;397: 928–940. doi:10.1016/S0140-6736(21)00458-X

7. Ozoh OB, Aderibigbe SA, Ayuk AC, Desalu OO, Oridota OE, Olufemi O, et al. The prevalence of asthma and allergic rhinitis in Nigeria: A nationwide survey among children, adolescents and adults. PLOS ONE. 2019;14: e0222281. doi:10.1371/journal.pone.0222281

8. Ozoh OB, Ayuk AC, Ukwaja KN, Desalu OO, Olufemi O, Aderibigbe SA, et al. Asthma management and control in Nigeria: the asthma insight and reality Nigeria (AIRNIG) study. Expert Review of Respiratory Medicine. 2019;13: 917–927. doi:10.1080/17476348.2019.1651201

9. Masekela R, Mortimer K, Nantanda R, Lesosky M, Meme H, Devereux G, et al. Asthma care in sub-Saharan Africa: Mind the gap! JPATS. 2022;3: 59–62. doi:10.25259/JPATS_12_2022

10. Ozoh OB, Eze JN, Garba BI, Ojo OO, Okorie E-M, Yiltok E, et al. Nationwide survey of the availability and affordability of asthma and COPD medicines in Nigeria. Trop Med Int Health. 2021;26: 54–65. doi:10.1111/tmi.13497

11. Plum C, Stolbrink M, Zurba L, Bissell K, Ozoh BO, Mortimer K. Availability of diagnostic services and essential medicines for non-communicable respiratory diseases in African countries. Int J Tuberc Lung Dis. 2021;25: 120–125. doi:10.5588/ijtld.20.0762

12. Stolbrink M, Thomson H, Hadfield RM, Ozoh OB, Nantanda R, Jayasooriya S, et al. The availability, cost, and affordability of essential medicines for asthma and COPD in lowincome and middle-income countries: a systematic review. The Lancet Global Health. 2022;10: e1423–e1442. doi:10.1016/S2214-109X(22)00330-8

13. Bissell K, Ellwood P, Ellwood E, Chiang C-Y, Marks GB, El Sony A, et al. Essential Medicines at the National Level: The Global Asthma Network’s Essential Asthma Medicines Survey 2014. Int J Environ Res Public Health. 2019;16: 605. doi:10.3390/ijerph16040605

14. Mallen. 2022 GINA Main Report. In: Global Initiative for Asthma - GINA [Internet]. [cited 8 Jul 2022]. Available: https://ginasthma.org/gina-reports/

15. Reddel HK, Bacharier LB, Bateman ED, Brightling CE, Brusselle GG, Buhl R, et al. Global Initiative for Asthma Strategy 2021: executive summary and rationale for key changes. Eur Respir J. 2022;59. doi:10.1183/13993003.02730-2021

16. Erhabor G, Abba A, Ozoh OB, Adeniyi B, Desalu OO, Falade AG, et al. Guideline for asthma management in Nigeria. Nigerian Thoracic Society; 2017.

17. Mortimer K, Reddel HK, Pitrez PM, Bateman ED. Asthma management in low- and middle-income countries: case for change. Eur Respir J. 2022; 2103179. doi:10.1183/13993003.03179-2021

18. The World Bank. Nigeria releases new report on poverty and inequality in country. In: World Bank [Internet]. 2020 [cited 7 Aug 2022]. Available: https://www.worldbank.org/en/programs/lsms/brief/nigeria-releases-new-report-on-poverty-and-inequality-in-country

19. Makinde OA, Sule A, Ayankogbe O, Boone D. Distribution of health facilities in Nigeria: Implications and options for Universal Health Coverage. Int J Health Plann Manage. 2018;33: e1179–e1192. doi:10.1002/hpm.2603

20. Technical working group (TWG). The National Strategic Health Development Plan Framework (2009-2015). 2009. Available: http://www.internationalhealthpartnership.net/fileadmin/uploads/ihp/Documents/Country_Pages/Nigeria/Nigeria%20National%20Strategic%20Health%20Development%20Plan%20Framework%202009-2015.pdf

21. Koce F, Randhawa G, Ochieng B. Understanding healthcare self-referral in Nigeria from the service users’ perspective: a qualitative study of Niger state. BMC Health Services Research. 2019;19: 209. doi:10.1186/s12913-019-4046-9

22. Tremblay S, Castiglione S, Audet L-A, Desmarais M, Horace M, Peláez S. Conducting Qualitative Research to Respond to COVID-19 Challenges: Reflections for the Present and Beyond. International Journal of Qualitative Methods. 2021;20: 16094069211009680. doi:10.1177/16094069211009679

23. Sandelowski M, Barroso J. Writing the proposal for a qualitative research methodology project. Qual Health Res. 2003;13: 781–820. doi:10.1177/1049732303013006003

24. Azevedo V, Carvalho M, Costa F, Mesquita S, Soares J, Teixeira F, et al. Interview transcription: conceptual issues, practical guidelines, and challenges. Rev Enf Ref. 2017;IV Série: 159–168. doi:10.12707/RIV17018

25. Ritchie J, Spencer L. Chapter 9: Qualitative data analysis for applied policy research. In: Burgess RG, Bryman A, editors. Analyzing qualitative data. London; New York: Routledge; 1994. pp. 173–194. Available: http://ezproxy.library.uq.edu.au/login?url=http://search.ebscohost.com/login.aspx?direct=true&db=nlebk&AN=72371&site=ehost-live

26. Gale NK, Heath G, Cameron E, Rashid S, Redwood S. Using the framework method for the analysis of qualitative data in multi-disciplinary health research. BMC Medical Research Methodology. 2013;13: 117. doi:10.1186/1471-2288-13-117

27. Creswell JW. Research design-qualitative, quantitative and mixed methods approach. 4th ed. London: SAGE publication; 2014. Available: https://scholar.google.com/scholar_lookup?title=Research%20design%E2%80%93qualitative%2C%20quantitative%20and%20mixed%20methods%20approaches&publication_year=2014&author=Creswell%2CJW

28. Stolbrink M, Chinouya MJ, Jayasooriya S, et al. Improving Access to Affordable Quality-Assured Inhaled Medicines in Low- and Middle-Income Countries | Request PDF. [cited 17 Aug 2022]. Available: https://www.researchgate.net/publication/361047940_Improving_Access_to_Affordable_Quality-Assured_Inhaled_Medicines_in_Low-_and_Middle-Income_Countries

29. Kavanagh J, Jackson DJ, Kent BD. Over- and under-diagnosis in asthma. Breathe. 2019;15: e20–e27. doi:10.1183/20734735.0362-2018

30. Joo MJ, Sharp LK, Au DH, Lee TA, Fitzgibbon ML. Use of Spirometry in the Diagnosis of COPD: A Qualitative Study in Primary Care. COPD: Journal of Chronic Obstructive Pulmonary Disease. 2013;10: 444–449. doi:10.3109/15412555.2013.766683

31. Akindele A, Daines L, Cavers D, Pinnock H, Sheikh A. Qualitative study of practices and challenges when making a diagnosis of asthma in primary care. npj Prim Care Respir Med. 2019;29: 1–7. doi:10.1038/s41533-019-0140-z

32. World Health Organisation. WHO package of essential noncommunicable (PEN) disease interventions for primary health care. 2020 [cited 2 Mar 2022]. Available: https://www.who.int/publications-detail-redirect/who-package-of-essential-noncommunicable-(pen)-disease-interventions-for-primary-health-care

33. Global Initiative for Asthma. Global Strategy for Asthma Management and Prevention. In: Global Initiative for Asthma - GINA [Internet]. 2020 [cited 21 Apr 2021]. Available: https://ginasthma.org/gina-reports/

34. Chukwuka C, Onyedum CC, Desalu OO, Ukwaja KN, Ezeudo C, Nwosu NI. Out-of-Pocket Costs of Asthma Follow-Up Care in Adults in a Sub-Saharan African Country. Journal of Respiratory Medicine. 2014;2014: 1–5. doi:10.1155/2014/768378

35. Li S, Odedina S, Agwai I, Ojengbede O, Huo D, Olopade OI. Traditional medicine usage among adult women in Ibadan, Nigeria: a cross-sectional study. BMC Complementary Medicine and Therapies. 2020;20: 93. doi:10.1186/s12906-020-02881-z

36. Galtung J. Violence, Peace, and Peace Research - Johan Galtung, 1969. 1969;6: 167–191. Available: https://journals.sagepub.com/doi/abs/10.1177/002234336900600301?journalCode=jpra

37. Egere U, Shayo E, Ntinginya N, Osman R, Noory B, Mpagama S, et al. Management of chronic lung diseases in Sudan and Tanzania: how ready are the country health systems? BMC Health Serv Res. 2021;21: 734. doi:10.1186/s12913-021-06759-9

38. Weller S. Using internet video calls in qualitative (longitudinal) interviews: some implications for rapport. 2017. doi:10.1080/13645579.2016.1269505

39. Santhosh L, Rojas JC, Lyons PG. Zooming into Focus Groups: Strategies for Qualitative Research in the Era of Social Distancing. ATS Scholar. 2021;2: 176–184. doi:10.34197/ats-scholar.2020-0127PS

